# Plasma Levels of Activin A and Follistatin-Like-3 as Biomarkers for Pulmonary Hypertension in Advanced Heart Failure

**DOI:** 10.1101/2025.03.06.25323530

**Authors:** Christopher Mackintosh, John Dreixler, Avery Tung, Ariel Mueller, Gene Kim, Sajid Shahul

## Abstract

**Introduction:** Pulmonary hypertension (pHTN) is a serious condition associated with increased mortality and morbidity, with limited early and non-invasive diagnostic tools. This study aimed to investigate the potential of plasma biomarkers Activin A and Follistatin-Like-3 (FSTL3) as indicators of disease severity in pHTN and right heart dysfunction.

**Methods:** We conducted a prospective cohort study of 120 patients with advanced heart failure undergoing right heart catheterization. Plasma levels of Activin and FSTL3 were measured at the time of the catheterization. Biomarker data was analyzed for correlation with values from right heart catheterization, including pulmonary artery pressure and right ventricular work.

**Results:** Both Activin A and FSTL3 levels were elevated in patients with pHTN compared to controls. Additionally, Activin A and FSTL3 were positively correlated with mean pulmonary artery pressure and right ventricular work, suggesting their potential as biomarkers for monitoring pHTN progression. In contrast, pro-BNP did not correlate with right ventricular work.

**Conclusion:** Results align with prior research indicating the importance of the activin signaling pathway in pHTN and support the hypothesis that measuring Activin and FSTL3 levels could offer a non-invasive method for diagnosing and monitoring disease progression in pHTN. Further studies are needed to validate these biomarkers in broader patient populations, which could ultimately enable earlier interventions and improve outcomes in pulmonary hypertension management.

## Introduction

Pulmonary hypertension (pHTN) describes a set of cardiopulmonary conditions that result in abnormally elevated pulmonary arterial pressure. Defined as a mean pulmonary artery pressure >20mmHg, pHTN may be caused by abnormal increases in pulmonary vascular resistance due to processes such as primary lung disease, pulmonary arterial hypertrophy, thromboembolic disease, or elevations in left atrial pressure due to cardiac conditions such as heart failure^**1**^. Regardless of underlying etiology, and despite advances in both medical management and lung transplantation, the 5-year mortality of this condition remains greater than 60 percent^**2**,**3**^. Diagnosis is made via right heart catheterization and is often delayed due to non-specific symptoms such as dyspnea or fatigue. The ability to screen for pulmonary hypertension and associated right ventricular dysfunction via non-invasive means such as blood testing could improve detection, allow more frequent monitoring for disease progression, and optimize treatment plans.

Activin and related proteins are members of the transforming growth factor beta (TGF-β) superfamily and play multiple roles in cell growth, inflammation, fibrosis, and other important processes^**4**^. In pulmonary arterial hypertension (PAH), activin pathway activity is increased, leading to increased downstream signaling of cell growth pathways including Smad2/3, MAPK, Wnt/ β-catenin, and PI3K/AKT. FSTL3 is a downstream target of ActRII signaling, and its expression is increased with pathway activation in cells. These pathways drive cell proliferation, inflammation, and fibrosis, which in turn lead to thickening of the pulmonary vasculature^**3**,**5**^. Increased activin activity has also been associated with pHTN in the setting of left heart failure^**6**,**7**^.

Emerging evidence suggests that blockade of activin signaling reverses the vascular remodeling that occurs during pHTN and improves cardiopulmonary function. In a 2023 phase III trial of patients with severe PAH, the small molecule activin inhibitor Sotatercept improved six-minute walk test performance at 24 weeks and reduced overall mortality compared to placebo^**8**^. These findings suggest that the activin pathway is an important driver of pHTN pathogenesis.

Activin A and its inhibitor Follistatin-Like-3 (FSTL3) have both been identified as potential biomarker candidates for PAH and pHTN. Elevated levels of both Activin and FSTL3 predict mortality in patients with pHTN^**3**^, raising the possibility that measuring serum levels of these biomarkers may allow clinicians to screen for or monitor the severity or progression of the disease. We thus hypothesized that serum levels of Activin A and FSTL3 would correlate with quantitative invasive measurements of pulmonary artery and right sided pressures. To test our hypothesis, we measured Activin A and FSTL3 levels in advanced heart failure patients immediately prior to their right cardiac catheterization procedures. These levels were then analyzed for correlation with measures of right ventricular performance.

## Methods

### Study Subjects

With appropriate IRB approval, we conducted a prospective cohort study of 120 patients with advanced heart failure referred to right heart catheterization. Patients were approached and consented to participate in the study on the day of their scheduled catheterization procedure. Patients were excluded from the study if they were younger than 18 years old, currently pregnant, had undergone heart transplantation, or had a left ventricular assist device in place. Consented patients agreed to the collection of one blood sample at the time of their procedure and review of their medical records including their most recent echocardiograms.

### Study Data

The electronic medical record was used to capture data about each subject, including demographic information, social history, NYHA class, co-morbidities, and concomitant medications. Blood samples were collected via venipuncture at the time of cardiac catheterization and analyzed via ELISA for plasma levels of Activin, FSTL3, and pro-BNP. Right heart catheterization was completed by routine institutional protocol and data for our research was collected from the clinical procedure report.

### Disease State Definitions

Presence of pHTN was determined quantitatively using data from cardiac catheterizations. Pulmonary hypertension was defined as all patients with a mean pulmonary artery pressure >20mmHg. Precapillary pulmonary hypertension was defined as mean pulmonary artery pressure >20mmHg and pulmonary vascular resistance >2 WU. Pulmonary hypertension due to left ventricular failure was defined as a pulmonary artery wedge pressure >15^**1**^. Patients meeting any of these sets of criteria were analyzed as the pulmonary hypertension group; those not meeting any of these criteria were analyzed as the control group.

### Data Analysis

Demographic information, disease characteristics, and baseline vital signs are summarized using descriptive statistics (**Table 1**). Because distributions of activin, FSTL3, and pro-BNP levels among study subjects were non-normal, we used Spearman’s correlations to analyze relationships between plasma biomarker levels and right heart catheterization values. Each of these three plasma biomarkers were evaluated for a correlation with right atrial and ventricular pressures, pulmonary artery and pulmonary capillary wedge pressure, and right ventricular work. Our pre-specified primary analysis involved the correlation between plasma activin and FSTL3 levels and right ventricular work.

**Table 1:** Baseline Characteristics.

| Table 1: Baseline Characteristics |  |
| --- | --- |
| Age, *years* | 59.31 +/- 12.58 |
| Height, *inches* | 68.49 +/- 4.30 |
| BMI, *value* | 27.88 (24.22, 34.02) |
| Gender |  |
| Male, *N(%)* | 77 (64.2%) |
| Female, *N(%)* | 43 (35.8%) |
| Smoking Status |  |
| Current Smoker, *N(%)* | 9 (7.5%) |
| Former Smoker, *N(%)* | 55 (45.8%) |
| Never Smoker, *N(%)* | 51 (42.5%) |
| Unknown, *N(%)* | 5 (4.2%) |
| Race |  |
| Black, *N(%)* | 61 (50.8%) |
| White, *N(%)* | 38 (31.7%) |
| Other/Unknown, *N(%)* | 16 (13.3%) |
| Asian, *N(%)* | 3 (2.5%) |
| Native American, *N(%)* | 1 (0.8%) |
| Latino, *N(%)* | 1 (0.8%) |
| Alcohol Use |  |
| No, *N(%)* | 73 (61.3%) |
| Yes, *N(%)* | 33 (27.7%) |
| Unknown, *N(%)* | 13 (10.9%) |
| NYHA Class |  |
| 1, *N(%)* | 3 (2.5%) |
| 2, *N(%)* | 29 (24.2%) |
| 3, *N(%)* | 69 (57.5%) |
| 4, *N(%)* | 19 (15.8%) |
| Pulmonary Hypertension, *N(%)* | 87 (72.5%) |
| LV Ejection Fraction, % | 29.8 ( 20.9, 47.7) |
| Heart Rate, *BPM* | 76 (66, 87) |
| Right Atrial Pressure, *mmHg* | 8 (5, 12) |
| Pulmonary Vascular Resistance, *Woods* | 1.88 (1.34, 3.09) |
| Pulmonary Capillary Wedge Pressure, *mmHg* | 16 (11.5, 24.5) |
| Pulmonary Artery Oxygen Saturation, % | 67 (60.12, 72.65) |
| Fick Cardiac Output, *L/min/m2* | 5.2 (4.28, 6.44) |
| Thermodilution Cardiac Output, *L/min/m2* | 4.3 (3.6, 5.4) |
| Systemic Vascular Resistance, *dynes\*sec/cm5* | 1248 (977, 1516) |
| Pulmonary Systolic Pressure, *mmHg* | 44 (30, 55) |
| Pulmonary Diastolic Pressure, *mmHg* | 20 (15, 28) |
| Mean Pulmonary Artery Pressure, *mmHg* | 28.33 (20, 37.33) |

## Results

### Baseline Demographics and Disease Characteristics

A total of 135 subjects were approached and consented for the study. 15 patients were later excluded from analyses due to a history of heart transplantation or having a left ventricular assist device in place. A total of 120 subjects were thus included in the final analysis. Baseline characteristics are described in **Table 1**. No patients were receiving inotropes, vasopressors, or mechanical support at the time of study. Median patient age was 59.31 (+/- 12.58) years. Patients were 64.2% male and were 50.1% black, 31.7% white, and 18.2 % other races.

### Cardiac Catheterization Data

The median left ventricular ejection fraction was 29.8% (20.9, 47.7). Median right heart catheter data were as follows: Right atrial pressure 8 mmHg (5, 12); pulmonary capillary wedge pressure 16 mmHg (11.5, 24.5); PA oxygen saturation 67% (60.12, 72.65); Fick cardiac output 5.2 L/min/m2 (4.28, 6.44); thermodilution cardiac output 4.3 L/min/m2 (3.6, 5.4); Mean Pulmonary Artery Pressure 28.33 mmHg (15, 28).

### Plasma Biomarkers

Patients with a diagnosis of pulmonary hypertension had higher levels of both plasma Activin (0.55 ng/mL vs. 0.43 ng/mL, p=0.04) and plasma FSTL3 (24.82 ng/mL vs 18.99 ng/mL, p=0.007) (see **Table 2**).

**Table 2.** Biomarker levels stratified by pHTN.

| 2a - Plasma FSTL3 Levels (ng/mL) p=0.0009 |  |  |  | 2b - Plasma Activin Levels (ng/mL) p=0.02 |  |  |  |
| --- | --- | --- | --- | --- | --- | --- | --- |
|  | 25<sup>th</sup> % | Median | 75<sup>th</sup> % |  | 25<sup>th</sup> % | Median | 75<sup>th</sup> % |
| **pHTN**<br>(n = 83) | **17.495** | **26.105** | **40.275** | **pHTN**<br>(n = 88) | **0.60375** | **0.803** | **1.157** |
| **No pHTN**<br>(n = 29) | **12.895** | **18.28** | **24.0415** | **No pHTN**<br>(n = 30) | **0.431625** | **0.6823** | **0.96925** |

Total N = 112; 8 patients with FSTL3 level below threshold for detection
Total N = 118; 8 patients with Activin level below threshold for detection

### Analyses of Catheter Data and Plasma Protein Data

Spearman’s Correlations were calculated to analyze the relationship between pressures recorded during right heart catheterization and plasma levels of activin, FSLT3, and pro-BNP. All correlation values are displayed in **Table 3**. Plasma levels of pro-BNP were not correlated with Right Atrial Pressure (p = 0.06), Right Ventricular Power (p = 0.12) or Right Ventricular Work (p = 0.53). but positively correlated with Mean Pulmonary Artery Pressure (p = 0.002),

**Table 3.**
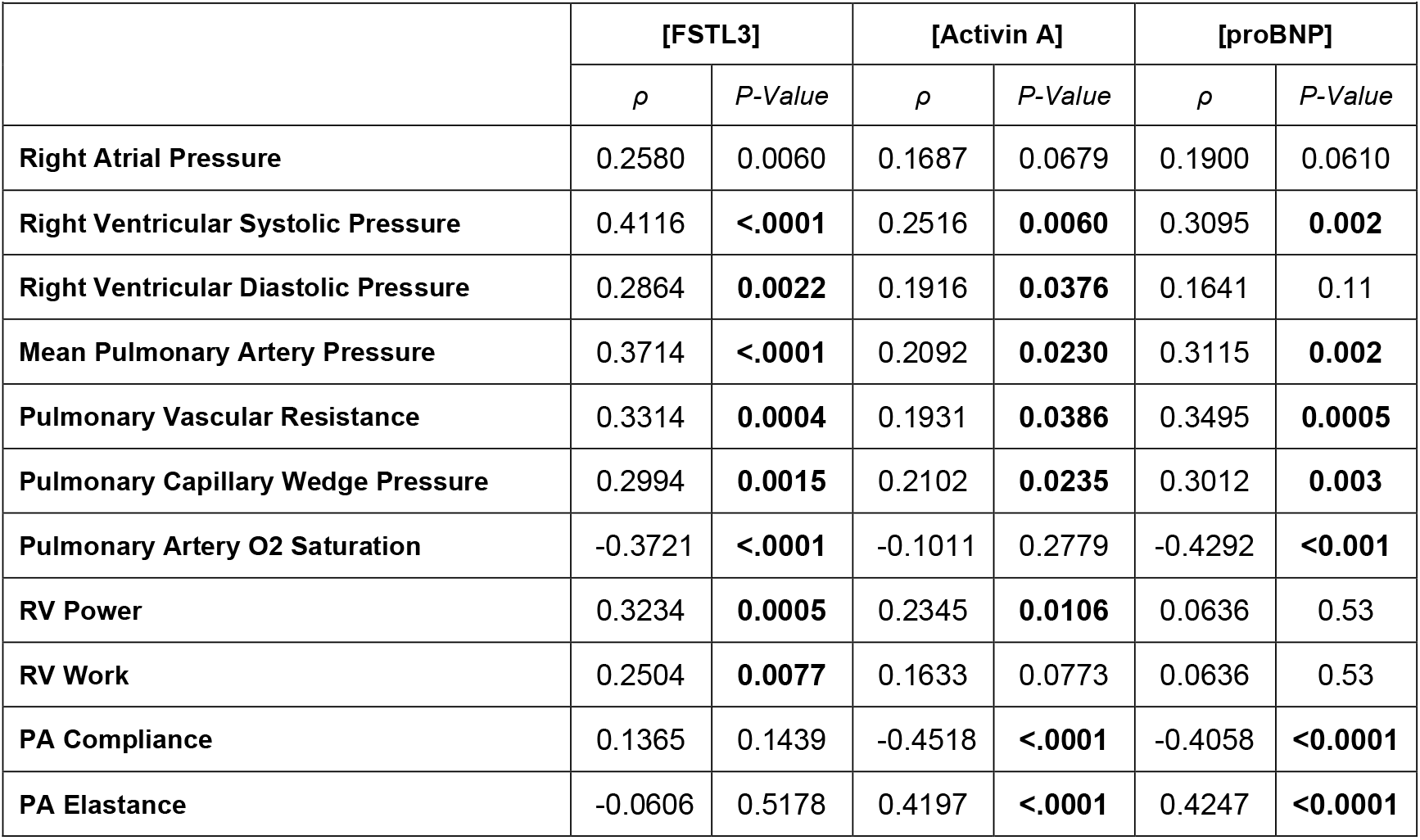
Pearson Correlation Coefficients.

Plasma activin levels were positively correlated with Mean Pulmonary Artery Pressure (p = 0.023) and Right Ventricular Power (p = 0.0106). Plasma FSTL3 was also positively correlated with Mean Pulmonary Artery Pressure (p <0.0001), Right Ventricular Work (0.0077), and Right Ventricular Power (p = 0.0005) (**Table 3**).

## Discussion

In our prospective observational trial of 120 patients who underwent concurrent measurement of right heart pressures and Activin A and FSTL3 levels, we found that plasma levels of these biomarkers correlated with increased pulmonary artery pressures and with greater right ventricular work and power. In addition, Activin and FSTL3 levels correlated with pulmonary vascular resistance and were higher in patients with pHTN than in those without. In contrast, plasma pro-BNP, a well-established marker for left ventricular failure, did not correlate with right ventricular strain. These findings together suggest that Activin and FSLT3 may be valuable plasma biomarkers for detecting pulmonary hypertension and right ventricular dysfunction.

These results align with prior research demonstrating the role of the activin pathway in pHTN and contribute further evidence that plasma levels of Activin and FSTL3 are elevated in patients with pHTN from diverse underlying causes. Previous studies have highlighted the over-expression of activin in pulmonary vascular tissues that drives cellular mechanisms and contribute to the pathogenesis of this disease^**3**^. The recent FDA approval of Sotatercept, a small molecule inhibitor of the activin pathway, further supports the biological and clinical relevance of activin related pathways in pulmonary hypertension^**8**^. Our study extends this body of work by correlating activin pathway proteins with disease severity as characterized by invasively measured right heart pressures.

Clinically, these data raise the possibility that the measurement of plasma levels of Activin and FSLT3 may provide a non-invasive method for clinicians to assess the progression of pulmonary hypertension. These biomarkers may serve as screening tools for diagnosis, complementary tools to guide clinical decision making, monitor disease progression, and determine the necessity of catheterization or other more invasive procedures. If validated in larger cohorts, Activin and FSTL3 assays could improve outcomes in pulmonary hypertension by better informing disease monitoring and enabling earlier interventions.

Our study has limitations. First, our sample size was small and limited to patients with advanced heart failure. It is possible that the correlations we observed may not be as strong in a more diverse patient population with a lower incidence of cardiorespiratory dysfunction. Additionally, our study examined a one-time cross section of heart failure patients and thus does not provide information regarding changes in biomarker levels with time and disease progression. We thus cannot say whether ongoing monitoring of Activin A and FSLT3 can detect meaningful changes in right heart pressures or function. Our data do however provide preliminary support for the investigation of plasma activin and FSTL3 values as diagnostic studies in these disease states.

In conclusion, our findings demonstrate that plasma levels of activin and FSTL3 are associated with increased right ventricular work in patients with known left ventricular heart failure. These biomarkers may be therefore valuable indicators of disease severity. Given the potential for non-invasive measurement, activin and FSTL3 could play a role in screening, monitoring disease progression, and guiding clinical decision-making in heart failure and pulmonary hypertension. Validation in larger, more diverse cohorts is essential to confirm clinical utility and to determine whether these studies can enable earlier detection and intervention, ultimately improving patient outcomes.

## Data Availability

Data included within manuscript tables.

